# Epidemiology of COVID-19 and effect of public health interventions, Chennai, India, March - October 2020

**DOI:** 10.1101/2021.04.21.21255852

**Authors:** M Jagadeesan, Parasuraman Ganeshkumar, Prabhdeep Kaur, Hemalatha Masanam Sriramulu, Manikandanesan Sakthivel, Polani Rubeshkumar, Mohankumar Raju, Lakshmidevi Murugesan, Raajkumar Ganapathi, Mahalakshmi Srinivasan, Aswini Sukumar, Kumaravel Ilangovan, Madhusudhan Reddy, Divyadarshini Shanmugam, Prakash Govindasamy, Manoj Murhekhar

**Author notes:** Corresponding author* Dr Prabhdeep Kaur, Scientist-E, ICMR- National Institute of Epidemiology, R-127, TNHB, Ayapakkam, Chennai- 600077, Tamil Nadu, India- 600 003. Joint first authors (M Jagadeesan, Parasuraman Ganeshkumar).

## Abstract

**Objectives:** To describe the public health strategies and their effect in controlling the COVID-19 pandemic from March to October 2020 in Chennai, India.

**Setting:** Chennai, a densely populated metropolitan city in Southern India, was one of the five cities which contributed to more than half of the COVID-19 cases in India.

**Participants:** We collected the de-identified line list of all the 192,450 COVID-19 case-patients reported from 17 March to 31 October 2020 in Chennai and their contacts for the analysis. We defined a COVID-19 case-patient based on the RT-PCR positive test in one of the Government approved labs.

**Outcome measures:** The primary outcomes of interest were incidence of COVID-19 per million population, case fatality ratio, deaths per million and the effective reproduction number (R_t_). We also analysed the indicators for surveillance, testing, contact tracing and isolation.

**Results:** Of the 192,450 RT-PCR confirmed COVID-19 case-patients reported in Chennai from 17 March-31 October 2020, 114,889 (60%) were males. The highest incidence was 41,064 per million population among the 61-80 years. The incidence peaked during June 2020 at 5239 per million and declined to 3,627 per million in October 2020. The city reported 3,543 deaths, with a case fatality ratio (CFR) of 1.8% and the crude death rate was 431 per million. When lockdown began, Rt was high (4.2) in March and fluctuated from April to June 2020. The R_t_ dropped below one by the first week of July and remained so until October 2020, even with the relaxation of restrictions

**Conclusion:** The combination of public health strategies controlled the COVID-19 epidemic in a large, densely populated city in India. We recommend continuing the interventions to prevent resurgence, even as vaccination is being rolled out.

**Strengths:**

- We did a comprehensive analysis of COVID-19 strategies and outcome in a large, densely populated metropolitan city in India.
- We documented that the community-centric public health strategies were feasible and effective in controlling the COVID-19 outbreak even in a large, thickly populated city
- The lessons learnt are relevant to similar settings in low-and middle-income countries. Given the ongoing multiple waves of COVID-19 and the difficulty in controlling the transmission, our experience and lessons learnt will be valuable for policymakers and scientific advisors globally

**Limitations:**

- We analysed the data available from the GCC database and not from the hospitals where patients with moderate to severe illness were admitted. Hence, we could not report the severity of illness among admitted patients.
- Second, the COVID-19 incidence might have been underestimated while testing was low during the early phase of the epidemic

## Introduction

COVID-19, which was reported from Wuhan, China, on 31 December 2019, has spread across the globe, affecting over 100 million individuals and causing over two million deaths by March 2021.[1] India reported its first case on 30 January 2020 and over 11 million confirmed cases and 158,856 deaths till 16 March, 2020.[1] Urban settings worldwide are major hubs for uncontained outbreaks,[2] posing challenges in controlling the transmission, and in mitigating economic and social hardship. Early in the pandemic, metropolitan areas faced the highest incidence of COVID-19.[3] Six months after the start of the pandemic, nearly 95% of COVID-19 cases reported globally were from urban areas.[2] In India, five cities accounted for nearly half of the COVID-19 cases reported till May 2020, with Chennai being a major contributor.[4] The predominance of inter-generational families, frequent socialisation of extended families and friends in the society, and more densely populated areas, challenged the public health actions taken to control the transmission.

To control the spread of COVID-19 in the country, the Government of India (GoI) imposed a country-wide complete lockdown on 25 March 2020. The lockdown was in place until 3 May 2020, following which several relaxations were given to sustain the economy. Despite the lockdown, Chennai, the capital city of the southern state of Tamil Nadu, faced challenges in the fight against the pandemic during the initial months. Chennai has an estimated population of over 8 million, with 31.5% residing in slum areas.[5] The city reported its first case of COVID-19 on 17 March 2020, and about 192,450 confirmed cases and 2452 deaths were reported until 31 October 2020.

The city health department had to overcome several challenges, including lack of adequate human resources for public health activities, large slum population, lack of awareness about the disease symptoms, and fear of being isolated/quarantined among the public. Closure of most private clinics and hospitals made passive surveillance difficult and led to overcrowding of patients at Government tertiary care hospitals for medical care and COVID-19 testing. Laboratories were overloaded, increasing the turnaround time and movement of people after the relaxation of restrictions posed challenges in contact tracing. Similar to Chennai, cities across the globe have been facing challenges in combating the COVID-19 pandemic.[6–8] The challenges include, but not limited to, high population density, high connectivity with other cities and urban areas, and unconventional interactions and communication leading to the rapid spread of false information.

Understanding these challenges, several health organisations had given additional attention and issued separate guidelines for mitigating COVID-19 transmission in urban settlements.[9–12] Besides policy level changes, the guidelines recommended mobilisation and capacity building of additional health workforce from different sources, community mobilisation and engagement, protecting and monitoring the vulnerable population, intensification of risk communication, establishing a call centre for coordination of public health response, setting up community-based testing sites, and data-driven decision making. Different strategies were adopted by several cities, according to their context, to control the pandemic. Describing such public health strategies, challenges in their implementation and the impact of these interventions would help policymakers make informed decisions during similar future situations in an urban setting. In this paper, we described the public health strategies and their effect in controlling the COVID-19 pandemic from March to October 2020 in Chennai, India.

## Methods

### Study Design and population

We analysed the COVID-19 data of Greater Chennai Corporation (GCC) from March to October 2020. Administratively, Chennai is divided into three regions (North, Central, and South), further divided into 15 administrative zones. Each Zone has 10-15 divisions, with a total of 200 divisions within the city.

Several densely populated areas and large wholesale markets for various commodities characterise Northern Chennai. The central region has one of the most crowded wholesale fruit/vegetable markets, commercial areas and office buildings. South Chennai is relatively less densely populated, with many offices of various software IT companies. The city has a well-structured public health system with a Zonal Health officer (ZHO) for each of the 15 zones responsible for the surveillance and response during epidemics. Each Zone has a dedicated health workforce for Medical and Public Health activities. Sanitary officers and sanitary inspectors are the frontline workers for all field-based public health activities.

### Description of Interventions

GCC implemented a comprehensive public health strategy for control of COVID-19, including surveillance, testing, contact tracing, isolation and quarantine (Supplementary table 1). The interventions were designed on the core principle of a community-centric patient-friendly approach. The surveillance and testing closer to home and field-based contact tracing in the streets with a cluster of cases based on epidemiological data analysis was the core strategy. Doctors and nurses examined symptomatic patients, collected samples in fever camps at 500+ locations daily, covering all streets in rotation. Another innovative strategy was identifying 3500 volunteers known as FOCUS (Friends of COVID persons Under Surveillance) volunteers, who were young adults from the same community.

**Table 1.** Incidence per million population and case fatality ratio of COVID-19 by age and gender, Chennai, India, March to October 2020.

| Characteristics |  | Mar'20 | Apr'20 | May'20 | Jun'20 | Jul'20 | Aug'20 | Sep'20 | Oct'20 | Overall |
| --- | --- | --- | --- | --- | --- | --- | --- | --- | --- | --- |
| **Incidence per million population** |  |  |  |  |  |  |  |  |  |  |
| **Sex** | **Male** | 4 | 147 | 2210 | 6751 | 6134 | 5447 | 4731 | 4799 | **30223** |
|  | **Female** | 2 | 86 | 1473 | 4640 | 4509 | 3762 | 3146 | 3086 | **20705** |
| **Age Group** | **≤20** | 1 | 46 | 636 | 1619 | 1923 | 1581 | 998 | 872 | **7677** |
|  | **21-40** | 3 | 139 | 2102 | 5929 | 5371 | 4696 | 3882 | 3831 | **25952** |
|  | **41-60** | 2 | 147 | 2566 | 8328 | 7433 | 6515 | 5843 | 6017 | **36852** |
|  | **61-80** | 11 | 120 | 2048 | 8935 | 8236 | 7039 | 7114 | 7562 | **41064** |
|  | **>80** | 12 | 110 | 1400 | 6596 | 6608 | 5757 | 6986 | 6207 | **33675** |
| **Total incidence** |  | **3** | **107** | **1694** | **5239** | **4894** | **4235** | **3623** | 3627 | **23422** |
| **Case Fatality Ratio (CFR) in %** |  |  |  |  |  |  |  |  |  |  |
| **Sex** | **Male** | 6.7 | 2.9 | 2.7 | 3.2 | 2.2 | 1.8 | 1.6 | **1.2** | **2.1** |
|  | **Female** | 0.0 | 0.0 | 2.2 | 1.9 | 1.5 | 1.3 | 1.1 | **0.8** | **1.4** |
| **Age group** | **≤20** | 0.0 | 0.0 | 0.1 | 0.1 | 0.1 | 0.0 | 0.0 | **0.0** | **0.1** |
|  | **21-40** | 0.0 | 0.5 | 0.4 | 0.4 | 0.3 | 0.1 | 0.1 | **0.0** | **0.2** |
|  | **41-60** | 0.0 | 4.2 | 3.0 | 2.5 | 1.6 | 1.3 | 1.0 | **0.2** | **1.6** |
|  | **61-80** | 14.3 | 8.9 | 12.2 | 9.8 | 7.6 | 6.4 | 4.5 | **1.2** | **6.9** |
|  | **>80** | 0.0 | 22.2 | 19.1 | 23.6 | 18.8 | 18.8 | 10.3 | **3.7** | **16.8** |
| **Total CFR** |  | **4.2** | **2.5** | **2.5** | **2.7** | **1.9** | **1.6** | **1.3** | **1.1** | **1.8** |

They visited the patients in home isolation with all precautions and ensured compliance with isolation and quarantine. They also assisted the families in purchasing groceries, medicines for their livelihood. They reported the visits and any violation of isolation and quarantine in an android application.

The state government implemented several Non-Pharmaceutical Interventions between March and October 2020 (Supplementary table 2). Between 5 February and 23 March 2020, international travel restrictions were in place. The Government of India (GoI) announced a complete lockdown across the country from 24 March to 14 April, further extending till 3 May 2020. All government and private institutions exempting essential services were closed during this period. All the transport services were suspended, excluding transportation of essential goods. All the educational and training institutes, worship places, and all functions/gathering were closed/suspended without any exemption. Wearing masks while moving out of the house was compulsory since mid-April. The international travellers who visited after 15 February 2020 were tested for COVID-19 with RT-PCR and were quarantined for 14 days at home. The Govt announced partial relaxations for offices from 4^th^ May 2020 onwards, and the partial travel resumed from 25 May 2020, with online registration and approval. From 1 June 2020, all private offices, showrooms, restaurants, tea shops, salons and taxis were permitted to operate with 50% capacity. Owing to a continuous increase in cases, the Government enforced a complete lockdown between 19 June and 5 July 2020. From July to October, the Government relaxed the restrictions in stages – increasing the maximum number of people who could attend marriage and funerals, increasing the number of domestic trains and flights, unrestricted inter-state and inter-district travel 14 August 2020 onwards. Starting from 1 September, the Government opened public parks and worship places for public use, allowed government and private offices to operate with 100% capacity, allowed hotels to accommodate guests, and started inter-district bus travel.

### Operational definitions

#### COVID-19 case

Any individual confirmed as infected with COVID-19 by testing the throat or nasal swab by RT-PCR technique. We used only RT-PCR as the diagnostic test to avoid misclassification, as the rapid antigen test had less sensitivity.

#### Contact of COVID-19 case

Any individual who had potential exposure to a COVID-19 case during the infectious period (3 days before to ten days after the date of onset of illness or the date of testing in case of asymptomatic cases). The potential exposures included staying in the same household, exposure within six feet/more than ten minutes, direct physical contact, providing care without using personal protective equipment, sharing food, travelling nearby for more than 15 minutes in a taxi or public or own transport, and sharing the same room such as meeting room and office room.

#### COVID Care Centre (CCC)

The centres which offered care only for patients that had been clinically assigned as mild or very mild cases.[13]

#### COVID-19 Hospital

The hospitals that offered comprehensive care primarily for those clinically assigned as severe.[13]

#### COVID-19 death

Any death due to a clinically compatible illness in an RT-PCR confirmed COVID-19 case unless there is an alternative cause of death that cannot be related to COVID diseases such as trauma without a period of complete recovery between illness and death.[14]

#### Home isolation criteria

The COVID-19 confirmed individuals who were asymptomatic or had very mild symptoms were eligible for home isolation after evaluation at screening centres, except for immunocompromised individuals, the elderly, and those with co-morbid conditions. All such individuals were considered discharged after ten days of symptom onset or date of testing and no fever for three days. After this period, the patients were advised to isolate and self-monitor their health further for seven days.[15]

#### Quarantine policy

All the contacts, as defined earlier, were quarantined at home for 14 days from the date of contact with a confirmed case.[16]

### Sources of data

We collected the line list of all the COVID-19 patients reported till 30 October 2020 from the GCC surveillance database. The line-list variables included the date of RT-PCR confirmatory report, age, sex, Zone of residence, hospitalisation status, and outcome. We retrieved daily testing data from the RT-PCR portal, a uniform nationalised data management platform for RT-PCR testing. We collected data regarding contact tracing, hospital occupancy, home isolation and quarantine data from an integrated online portal developed exclusively for COVID-19 response. We removed personal identifiers before data extraction. We obtained approval from the Institutional Ethics Committee for the analysis of anonymised data.

### Outcomes of interest

The study outcomes included incidence per million, case fatality ratio, death rate and the effective reproduction number. (a) Incidence of COVID-19 measured as the number of RT-PCR confirmed cases per million population by age, gender and Zone. (b) The case-fatality ratio was estimated as the number of deaths divided by the number of cases. (c) The death rate was defined as the number of COVID-19 deaths per million population. (d) The effective reproduction number R_t_ was defined as the number of cases emerging from one index case in the population.

### Data analysis

We described RT-PCR confirmed COVID-19 case-patients and COVID-19 deaths by time, place and person characteristics. We described time distribution by plotting the epidemic curve using the date of reporting. We estimated COVID-19 incidence, CFR and the death rate by age and gender. To describe the spread of COVID-19 across the geographical zones over time, we used qGIS software version to plot maps with the incidence per million population. We used Friedman’s test to test the difference in incidence across the age-groups and gender over time. We assessed the age and gender difference in CFR using the chi-square test. We used the “COVID-19 Estimator” developed by WHO-Pan American Health Organization (PAHO) office to estimate effective reproduction number (R_t_) with 95% credible interval.[17] The estimator was developed using the “EpiEstim” R package.[18] The estimator used the number of COVID-19 cases reported daily to estimate R_t_, assuming parametric serial interval distribution based on a mean (SD) of 4.8 (2.3) days.

We analysed the surveillance indicators, including the number of outreach camps and the proportion of all samples collected in camps or sample collection centres. We analysed the average number of tests done per million population per day, and test positivity %, defined as the number of tested positive among tested. The complete data on testing was available only from May 2020, when the entry of sample collection data in a centralised data entry portal and mobile application was made mandatory for sample collection centres and laboratories. We assessed the contact tracing activity by the estimating median number of contacts traced per COVID-19 case from July 2020, when contact tracing data was integrated into the online portal developed exclusively for COVID-19 response. We categorised the contacts as home (household) contacts and extended (non-household) contacts. We calculated the average bed occupancy rate for a month by taking an average of the daily bed occupancy rate.

### Patient and public involvement

We collected the de-identified data directly from the GCC surveillance database. The study team did not access the information such as patient identifiers or any other personal data. We did not involve the patients or public directly in the study design, outcome measures, data analysis, or interpretation of the results.

## Results

### Descriptive analysis of cases and deaths

Chennai reported 192,450 RT-PCR confirmed COVID-19 case-patients from 17 March 2020 until 31 October 2020, of which 114,889 (60%) were males, 74,635 (39%) belonged to 21-40 years and 66,616 (35%) to 41-60 years (Supplementary Table 3). The daily number of cases increased during May and peaked during June, after which it gradually declined (Figure 1).

**Figure 1.**
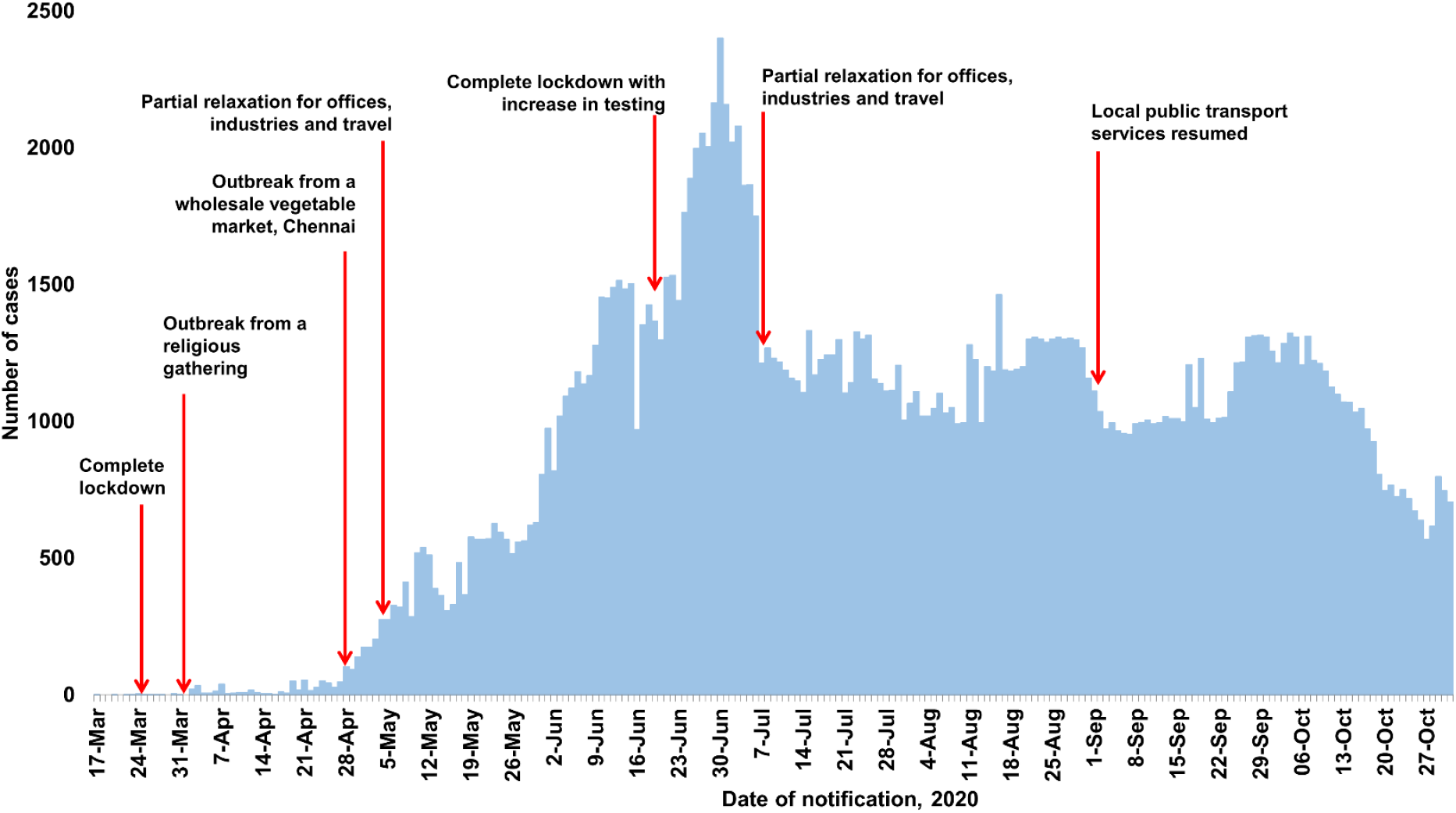
Distribution of RT-PCR confirmed COVID-19 cases by date of notification, Chennai, India, March to October 2020

As of 31 October 2020, the incidence of COVID-19 per million population was 23,422, with males (30,223 per million) reporting a higher incidence than females (20,705 per million) (Table 1). Among various age groups, the 61 – 80 years age group had the highest incidence (41,064 per million) and the less than 20 years age group had the lowest incidence.

The city reported 3,543 deaths during the same period. Among them, 2,452 (69%) were males, and 1,852 (52%) were of age group 61 – 80 years (Supplementary Table 3). Overall case fatality ratio (CFR) was 1.8% as of 31 October 2020 (Table 1). CFR was higher among males than females (2.1% and 1.4%, p-value<0.001). CFR increased with age (p-value<0.001) and was highest among people above 80 years (16.8%), followed by 61-80 years (6.9%).

The overall crude death rate of COVID-19 in Chennai city was 431 per million between March and October 2020. In line with CFR, the death rate was also higher among males (645 per million) than females (291 per million). The age-specific death rate increased with age increase, the highest being reported among people over 80 years (5635 per million), followed by 61-80 years age group (2817 per million).

### Effect of public health interventions

#### From initial case till May 3, 2020

Owing to intense nationwide-lockdown measures since March 24, 2020, the incidence of COVID-19 was 3 per million during March 2020 and 107 per million during April 2020. All the zones reported an incidence of less than 2300 per million during this period (Figure 2). The incidence was comparable in the 21-40, 41-60, and 61-80 years age group. CFR was high (4.2%) during March 2020 and reduced to 2.5% in April 2020. The effective reproduction number (R_t_) was high (4.2) during March due to a super spreader event from a religious gathering,[19] and reduced during April 2020 to reach 2.1 (Figure 3). At the end of April, there was a sudden increase in transmission due to another super spreader event from a wholesale market (Figure 1).[20] This increased the R_t_ to 2.4 towards the end of April (Figure 3).

**Figure 2.**
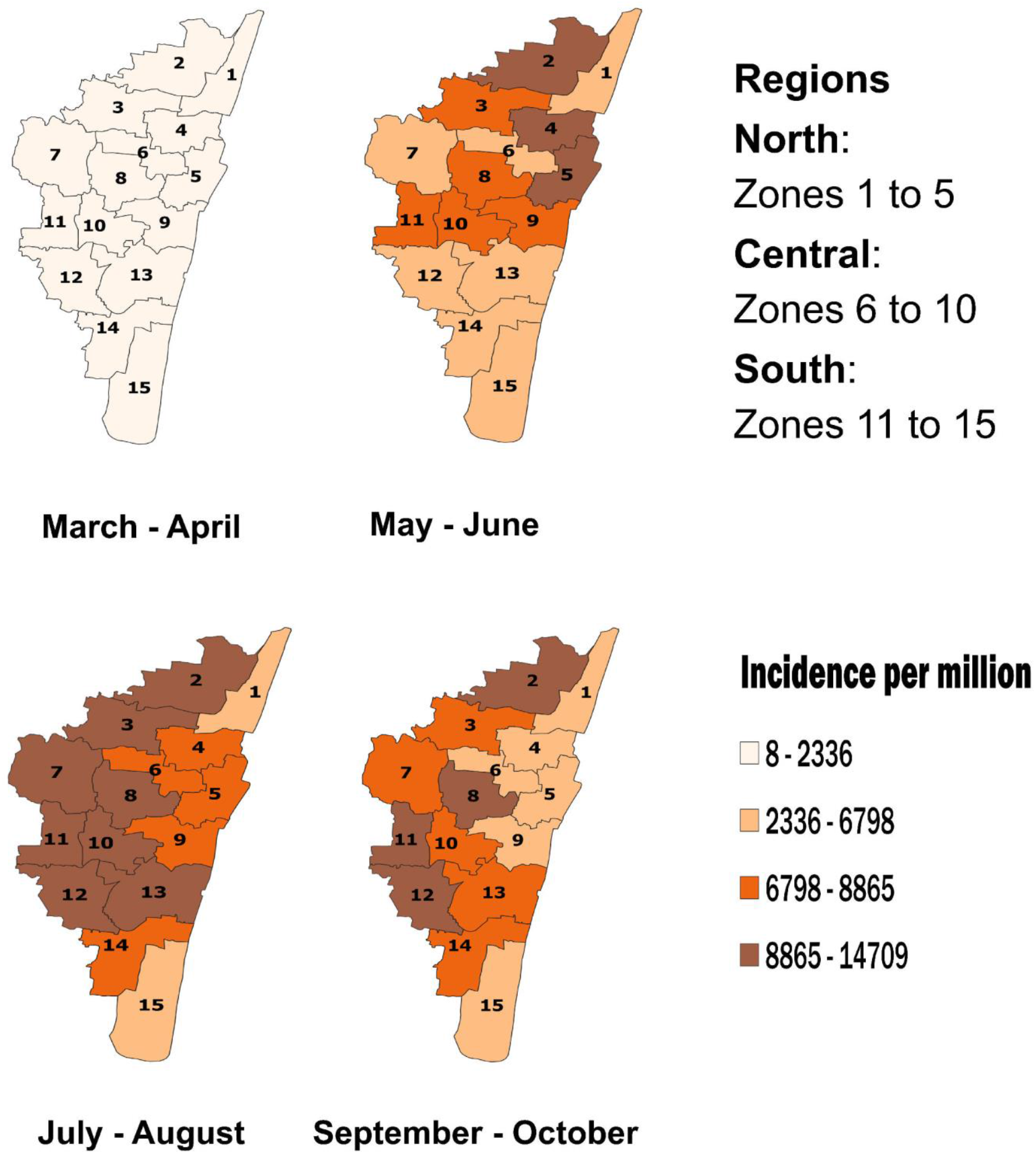
Incidence per million population by zone and month, Chennai, India, March to October 2020

**Figure 3.**
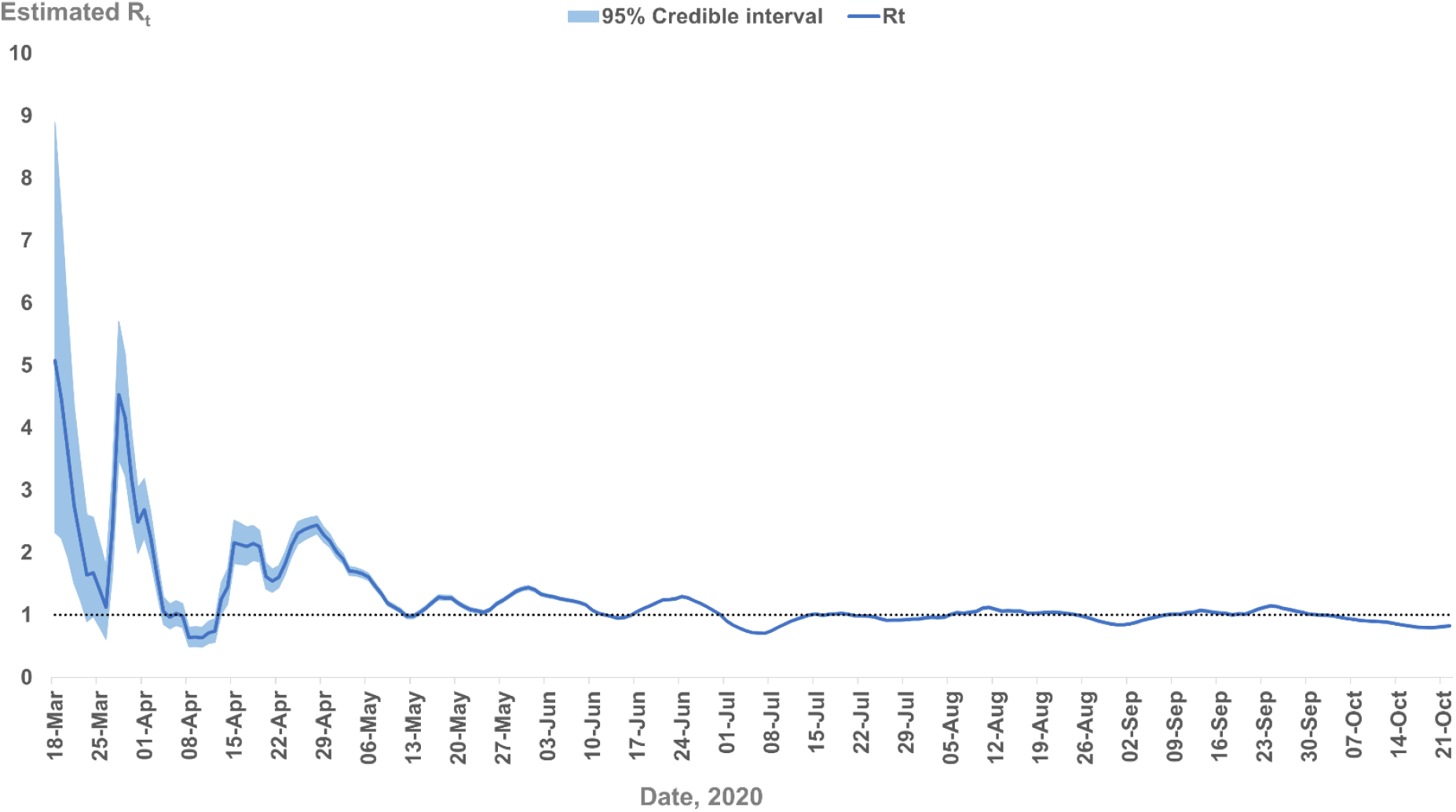
The Effective Reproduction Number (Rt) Estimates based on Rt-PCR Confirmed COVID-19 Cases in Chennai, India, March – October, 2020

#### May 4 to June 18, 2020

Following partial relaxation for domestic travel and workplaces and a super spreader event from the wholesale market,[20] the incidence increased to 1694 per million in May 2020. The CFR maintained at 2.5% during May, and about 4% of the active COVID-19 patients were isolated at home. After the wholesale market closure, the R_t_ reached 1.4 at the end of May 2020 (Figure 3). To increase testing, the GCC started outreach fever camps (17 camps per day) in May and established 35 sample collection centres closer to the community for wider testing and easy accessibility. On average, GCC conducted about 417 tests per million per day during May, with the help of 18 RT-PCR labs (six public and 12 private). The average daily test positivity was 12% during May (Figure 4). However, the incidence continued to increase during the initial days of June 2020, with an increase in CFR to 2.7%. The test positivity increased to 25% during June (Figure 4). The incidence was remarkably higher among adults above 40 years and in three zones in the northern region (Figure 2). The proportion of active patients under home isolation increased to 22%, and the average bed occupancy rate (total COVID-19 beds – 10469) in public and private sector went up to 69% and 46%, respectively. The oxygen bed occupancy was 70%, and ICU occupancy ranged from 41% in public to 68% in the private sector.

**Figure 4.**
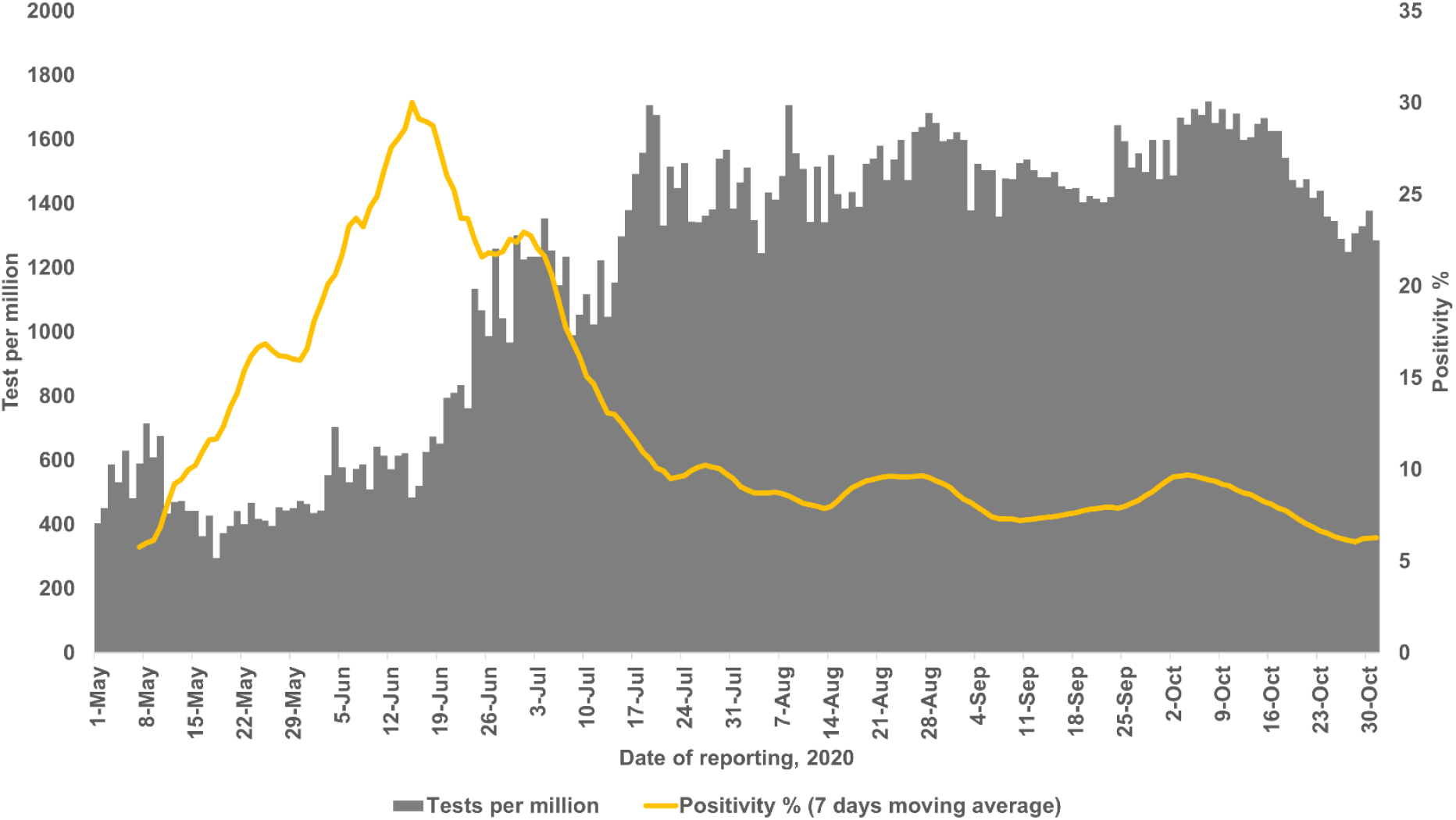
Test per million and positivity % by date of reporting, Greater Chennai Corporation, May to October 2020

#### June 18 to July 5 2020

Given the continued rise in incidence and bed occupancy, the state authorities implemented another complete lockdown to reduce the transmission. During this period, the number of outreach fever camps per day increased from 17 to 326 and the number of RT-PCR labs from 18 to 26. This led to an increase in the average number of tests done per million per day to 553, and the daily reported cases peaked on June 30, 2020 (Figure 1), with an incidence of 5239 per million. The test positivity declined to 21% at the end of the lockdown period (Figure 4). The R_t_ increased to a maximum of 1.3 on June 24, 2020, and declined thereafter to below 1.0 in the first week of July (Figure 3).

#### July 6 to October 2020

After the relaxation of restrictions similar to the pre-lockdown period, surveillance was intensified with 497 outreach fever camps per day in July and maintained over 400 until October. The number of sample collection centres increased from 35 to 56 in August. A total of 52 labs (13 public and 39 private) enabled average daily testing of over 800 per million and a decline in the test positivity to 8% (Figure 4). More than half of the total samples collected were from the outreach camps and dedicated sample collection centres. The city traced and quarantined over a million contacts between July and October. In July, the ratio of extended to home contacts was two and increased to three between August and October. The median (IQR) number of contacts traced per case detected in July was 11 (6-17). The median increased to 13 (8-17) in August 2020 and remained at the same level until October 2020.

The incidence gradually declined to 3,627 per million in October. From July until October, the incidence was highest among the 61-80 years age group, followed by the 41-60 years age group. The gender (p-value – 0.04) and age groups (p-value <0.001) significantly differed in terms of change of incidence from March to October 2020.

The incidence increased above 8800 per million in eight zones in the northern, central and southern regions during July-August. By October, only three zones in the central and southern region and one in the northern region reported incidence above 8800 per million. Of 13 zones, which had incidence above 6800 per million during July-August, four reported a decline during September-October (Figure 2). The R_t_ maintained at lower levels after the first week of July, though it fluctuated around 1.0 and reached 0.8 towards the end of October (Figure 3).

The bed capacity (occupancy%) in COVID-19 hospitals increased to over 14000 (42%) by July and declined to nearly 10000 (31%) by the end of October. This change was mainly due to a 50% decline in the private sector owing to low bed occupancy. In CCCs, the numbers maintained over 15000 throughout the period with a decline in occupancy from 33% in June to 2.8% in October. The oxygen bed occupancy declined to below 20% by October. The ICU bed occupancy reduced to 24% in public and 43% in the private sector. The CFR was gradually brought down to 1.1 by October (Table 1).

## Discussion

We documented the COVID-19 response in a large metropolitan city in south India. Our analysis suggested that the combination of interventions slowed the transmission and resulted in the decline of cases despite easing the restrictions. Unlike the effectiveness of lockdown measures in China and European countries, which led to a decline in cases,[21–26] poor compliance to restrictions in highly congested urban slums required adaptation of strategies tailored to the local setting. Therefore, we designed and implemented a community-centric public health strategy to improve early detection of clusters, access to free testing closer to the residence and free hospital-based care. This people-centric response was possible due to strong political will, good governance, extensive public sector hospital and laboratory infrastructure and dedicated human resources. Besides, a dedicated state-level procurement agency enabled rapid and timely procurement of PPE, consumables, laboratory equipment, and other hospital-based requirements.

We responded to the pandemic by rapid mobilisation of resources to implement a community-centric strategy, despite a resource-limited urban public health care system. Unlike rural India, which has a more structured public health care system with an extensive network of community health workers,[27] urban health systems are more fragmented, with many people seeking care in the private sector facilities.[28] Hence, the control strategy required the mobilisation of additional resources to implement various public health strategies such as outreach camps with sample collection, home isolation monitoring and support and establishing adequate institutional isolation/ quarantine facilities in addition to hospital-based care. The program managers recognised the need for an additional workforce early in the outbreak. They mobilised a vast network of paid volunteers for field activities and medical and public health personnel for clinical and managerial roles. The paid volunteers were young boys and girls from the same community. Hence they could easily connect with families in home isolation. In addition to monitoring compliance with the isolation, they also supported the family in procuring groceries, medicines and facilitated linkages with the health system. These strategies improved the acceptance of public health interventions at the community level. Tamil Nadu has a well-structured well-staffed state-level public health department with a highly trained workforce catering predominantly to the rural population. This workforce was mobilised to support various managerial and field level activities such as outreach camps, contact tracing, sample collection, data analysis, clinical care and strategic planning. The rapid scale-up of test-trace-isolate strategies with additional resources led to control of the outbreak. A similar community-based strategy helped Dharavi, India’s largest slum, reduce the spread of the infection.[29] Increased testing closer to home effectively reduced the cases and test positivity, indicating a slowdown in the transmission. A similar pattern was observed in other places such as New York, South Korea, New Zealand in the early phase of the epidemic.[30–32]

The data-driven decision making anchored the public health response to the pandemic in our setting. We established an integrated data management system that pooled the information from laboratories, hospitals, and field-level staff. The real-time data enabled daily and weekly analysis of cases up to the street level to identify the areas where surveillance and testing needed to be intensified. The regular analysis documented the gradual shift in higher incidence from northern Chennai to central and southern regions as the pandemic progressed. Accordingly, the program managers could rapidly shift the resources to control newly emerging hotspots. We learned from other cities such as New York and Singapore, which had data-driven approaches evident from the dashboards available on their websites.[33,34] We reviewed the key indicators recommended in the literature and adapted the relevant indicators to our setting.[35–37] In our experience, test positivity, incidence by geographic areas, case fatality ratio and % bed occupancy, were extremely useful in assessing the situation and adapting the strategies.

We observed unique epidemiological characteristics such as higher incidence in the older age group as the epidemic progressed. The incidence remained low among the <20 years age group irrespective of the month. Low incidence was due to the closure of educational institutions throughout the period. To begin with, the incidence was comparable across the age groups. However, from June onwards, as the cases increased rapidly, the elderly (>60 years) experienced a high burden. The pattern contrasts with the pattern observed in the United States and Europe, which reported a higher incidence in the younger age group as the epidemic progressed.[38] The multi-generational joint family system or frequent interactions of the elderly with other age groups at the societal level possibly facilitated the transmission. Given the asymptomatic or mild symptomatic status of younger people with COVID-19, they might have passed the infection to the elderly without their knowledge. The elderly were also more likely to have a symptomatic infection and hence higher chances of being tested and diagnosed.

We observed a decline in the case fatality ratio with time. The decline in CFR was possibly due to various factors such as early identification of hypoxia in outreach camps, improved management protocols, and awareness among patients regarding the need to seek care for COVID-19 like symptoms. However, the CFR continued to be high in older age groups, as witnessed in other cities and countries.[39–42] Despite the differences in overall CFR across the countries, age-specific CFR was considerably higher among individuals >60 years, as witnessed in this study and in China and Italy.[40] Individuals with age >80 years succumbed to death more than other age groups. In Chennai, the CFR in this age group was 16.8%, comparable to China (14.8%) and Italy (17.9%).

During the eight months (March to October 2020), Chennai reported 431 COVID-19 deaths per million. During 2019, the overall mortality rate was 769 per million population in Tamil Nadu state, India.[43] Due to communicable, maternal, neonatal, and nutritional diseases, the mortality rate was 113 per million and all the respiratory infections together reportedly caused 49 deaths per million.[43] Despite the control measures, reported COVID-19 deaths were many-fold higher than the expected mortality due to infectious diseases.

## Strengths and limitations

One limitation was that we analysed the data available from the GCC database and not from the hospitals where patients with moderate to severe illness were admitted. Hence, we could not report the severity of illness among admitted patients. Second, the COVID-19 incidence might have been underestimated while testing was low in the early phase. The strength of our study was the comprehensive analysis of COVID-19 strategies and outcome in a large, densely populated metropolitan city in India. The lessons learnt are relevant to similar settings in low-and middle-income countries.

## Conclusion

We conclude that the community-centric public health strategies controlled the COVID-19 outbreak in a large, thickly populated city in India. These efforts led to control in the short term. However, COVID-19 is an opportunity to strengthen the public health and primary health care system for the urban poor.

Since only one-third of the population antibodies for COVID-19 in October,[44] a large proportion of the population remained susceptible. Therefore, there is a resurgence risk due to new variants,[45] and poor compliance to COVID-19 appropriate behaviours. We need to sustain the public health surveillance, expand the public health workforce, educate communities regarding COVID-19 appropriate behaviours and sustain the test-trace-isolate strategies to prevent the resurgence of COVID-19 in Chennai.

## Supporting information

Strobe Checklist

## Data Availability

Data are available upon reasonable request.

## Acknowledgements

We thank all the Greater Chennai Corporation health staff, including Zonal Health Officers, doctors, nurses, field staff, volunteers who implemented COVID19 control interventions. We acknowledge the contribution of MPH scholars of ICMR-NIE, Chennai, who provided technical support to the Greater Chennai Corporation to implement public health interventions. We also thank all the departments of Greater Chennai Corporation and the Health and Family Welfare Department, Government of Tamil Nadu, who supported the COVID-19 response for Chennai.

## Contributorship statement

JM and PG contributed equally to the paper as joint first authors. JM designed the study, helped in data acquisition and revised the manuscript. PG, PK and MS designed the study, planned data analysis and wrote the first draft of the paper. AS helped in data acquisition, analysed the data and reviewed the manuscript. PR, MR and KE contributed to data analysis, interpretation of data and gave critical comments in finalising the manuscript. HM, LM, RG and MS managed data, helped interpret the results and revise the manuscript. MM interpreted the results, helped in drafting the manuscript and gave critical comments to finalise the manuscript. MR, DS and PG managed data, implemented the strategies and reviewed the manuscript. All authors approved the final manuscript. PK is the guarantor. The corresponding author attests that all listed authors meet authorship criteria and that no others meeting the criteria have been omitted. The views presented here are those of the authors and should not be attributed to ICMR-NIE.

## Ethical approval

The de-identified data were extracted from the GCC surveillance database after strict data protection protocols agreed between GCC and ICMR-NIE. We obtained approval from the Institutional Ethics Committee of ICMR-NIE for the study, with the project ID NIE/IHEC/202004-07. Since we did not interview the patients and used only the de-identified data from the database for the study, informed consent was not required.

## Provenance and peer review

Not commissioned; externally peer reviewed

## Data sharing statement

Data are available upon reasonable request.

## Dissemination declaration

Dissemination of the results to the patients is not applicable. We intend to disseminate the results to the policy makers

## Trial registration

Not applicable

