## Supplementary Table 1 for "Epidemiology of COVID-19 and effect of public health interventions, Chennai, India, March - October 2020"

**Supplementary table 1. Public health interventions implemented by Greater Chennai Corporation to control COVID-19 in Chennai, India, 2020**

| **Strategy** | **Interventions** |
| --- | --- |
| **Surveillance** | Passive surveillance in all Government tertiary care facilities, COVID-19 sample collection centres and private hospitals approved for COVID-19 care |
|  | Outreach fever camps in 500+ locations every day from May 2020 onwards with a team of doctor, nurse and lab technician in full PPE to evaluate the symptomatic individuals and contacts of confirmed cases. All the individuals who attended the camps were evaluated for hypoxia using a finger pulse oximeter. |
|  | 10,000+ volunteers (18 to 25 year-old) did house to house visits to identify the symptomatic individuals and referred them to fever camps or testing centres |
| **RT PCR Testing** | Sample collection was done in 42 new sample collection centres, 10 mobile collection units, 4 Government hospitals, besides private hospitals and laboratories. All individuals who got tested were evaluated for hypoxia using a finger pulse oximeter. |
|  | Testing done in 13 Government and 39 private labs with total capacity to process 15000+ samples per day |
| **Contact tracing** | Dedicated call centre with 200+ trained contact tracers |
|  | Sanitary inspectors visited the households of COVID-19 cases for contact tracing |
| **Triaging of COVID-19 confirmed cases** | 15 triaging centres established to screen COVID-19 patient, patients advised home isolation or referred to hospital-based on age, severity, co-morbidities and laboratory tests |
| **Home isolation/ Quarantine** | Phone-based monitoring of patients in home isolation by a dedicated team of doctors and paramedics and facilitated hospital transfer based on the clinical condition if required. |
|  | Home quarantine and isolation monitoring by paid FOCUS (Friends of COVID persons Under surveillance) volunteers who visited the COVID-19 patients and contacts to monitor health status/ compliance and provided support for procuring groceries, medicines or any other needs. |
| **Hospital-based care** | 80 ambulances, 64 mobile medical units, 60 mini-buses, 20 special services vehicles (102 services), and 65 twelve-seater vehicles to transport the patients |
|  | Government COVID care centre beds, tertiary and secondary care hospitals with oxygen beds and ICU beds |
| **Integrated data management system and data analysis** | An integrated data management system used at all levels to capture data from various levels from sanitary inspector to the control room. The data elements included a line list of cases with address, line list of contacts, the outcome of each case and bed occupancy status. |
|  | Rigorous data analysis up to street level to map the hotspots for planning fever camps, aggressive testing and intensifying non-pharmacological interventions. |
| **Mandatory usage of face mask** | Government advised the usage of face mask by the people during interaction with others since April 2020. The usage of the face mask was made mandatory while in public places since mid-April 2020. |

**Supplementary table 2. Various activities allowed during different periods of lockdown in Chennai, India, 2020**

| **From** | | **To** | **International travel** | **Domestic road travel** | **Domestic train travel** | | | **Domestic air travel** | **Local Public transport** | **Educational institutions** | **Offices** | **Industries** | | | **Worship places** | | **In-house dining** | **Gathering** | **Hotels and recreation clubs** | **Remarks** |
| --- | --- | --- | --- | --- | --- | --- | --- | --- | --- | --- | --- | --- | --- | --- | --- | --- | --- | --- | --- | --- |
| 05-Feb | | 23-Mar | **Ƥ** | **✓** | **✓** | | | **✓** | **✓** | **✓** | **✓** | **✓** | | | **✓** | | **✓** | **✓** | **✓** | Visas cancelled for those from selected countries |
| 24-Mar | | 03-May | **🞬** | **🞬** | **🞬** | | | **🞬** | **🞬** | **🞬** | **🞬** | **🞬** | | | **🞬** | | **🞬** | **🞬** | **🞬** | Except for essential services. Funeral - 20 persons allowed |
| 04-May | | 31-May | **Ƥ** | **Ƥ** | **Ƥ** | | | **Ƥ** | **🞬** | **🞬** | **Ƥ** | **Ƥ** | | | **🞬** | | **🞬** | **🞬** | **🞬** | Government offices - 50% capacity, IT offices - 10%, and industries - 25%. All stand-alone shops could operate except air-conditioned establishments, salons, spa, and beauty parlours. Funeral & marriage - 20 persons allowed. Repatriation international flights started under Vande Bharat Mission |
| 01-Jun | | 18-Jun | **Ƥ** | **Ƥ** | **Ƥ** | | | **Ƥ** | **Ƥ** | **🞬** | **Ƥ** | **Ƥ** | | | **🞬** | | **Ƥ** | **🞬** | **🞬** | All private offices, showrooms, restaurants, tea shops, barbershops and beauty parlours permitted to operate with 50% capacity without air conditioning. Public transport - 50% capacity, only within Greater Chennai Corporation limits. Funeral & Marriage - 50 persons allowed |
| 19-Jun | | 05-Jul | **Ƥ** | **🞬** | **Ƥ** | | | **Ƥ** | **🞬** | **🞬** | **Ƥ** | **Ƥ** | | | **🞬** | | **🞬** | **🞬** | **🞬** | Owing to a continuous increase in cases, the Government enforced a complete lockdown with minimal permissions. Government offices and banks could operate at 33% capacity. Inter-state and inter-district travel were restricted only to essential travel through pre-travel online registration and approval. |
| 06-Jul | | 31-Jul | **Ƥ** | **Ƥ** | **Ƥ** | | | **Ƥ** | **Ƥ** | **🞬** | **Ƥ** | **Ƥ** | | | **🞬** | | **Ƥ** | **🞬** | **🞬** | Same as pre-lock down period (1 June, 2020, to 18 June, 2020), with the exception to open sports complexes and stadia with no spectators. Interstate trains and flights were allowed. Funeral - 20 persons & Marriage - 50 persons allowed |
| 01-Aug | | 31-Aug | **Ƥ** | **Ƥ** | **Ƥ** | | | **Ƥ** | **Ƥ** | **🞬** | **Ƥ** | **Ƥ** | | | **🞬** | | **Ƥ** | **🞬** | **🞬** | Private offices, industries, restaurants and tea shops permitted to operate at 50% capacity. Independence Day celebrated with strict social distancing and mask compliance in Government premises. Since 14 August, inter-district and inter-state travel resumed with no restrictions on the movement of people. Funeral - 20 persons & Marriage - 50 persons allowed |
| 01-Sep | | 30-Sep | **Ƥ** | **✓** | **✓** | | | **✓** | **✓** | **🞬** | **✓** | **✓** | | | **✓** | | **✓** | **🞬** | **✓** | Public and private bus transport and metro rail services allowed between districts and within the city, but not between states. All except educational institutions were permitted to open with 100% capacity. Funeral - 20 persons & Marriage - 50 persons allowed |
| 01-Oct | | 31-Oct | **Ƥ** | **✓** | **✓** | | | **✓** | **✓** | **🞬** | **✓** | **✓** | | | **✓** | | **✓** | **🞬** | **✓** |  |
| **🞬** | Not allowed | | | | | **Ƥ** | Allowed partially | | | | | |  | **✓** | | Allowed with 100% capacity | | | | |

**Supplementary Table 3. Distribution of RT-PCR confirmed COVID-19 cases and deaths by age and gender, Chennai, India, March to October 2020**

| **Characteristics** | | **Mar'20** | | **Apr'20** | | **May'20** | | **Jun'20** | | **Jul'20** | | **Aug'20** | | **Sep'20** | | **Oct'20** | | **Overall** | |
| --- | --- | --- | --- | --- | --- | --- | --- | --- | --- | --- | --- | --- | --- | --- | --- | --- | --- | --- | --- |
|  |  | **#** | **%** | **#** | **%** | **#** | **%** | **#** | **%** | **#** | **%** | **#** | **%** | **#** | **%** | **#** | **%** | **#** | **%** |
| **COVID-19 cases** | |  |  |  |  |  |  |  |  |  |  |  |  |  |  |  |  |  |  |
| **Sex** | **Female** | 9 | 38 | 322 | 37 | 5518 | 40 | 17381 | 40 | 16892 | 42 | 14093 | 40 | 11783 | 40 | 11561 | 39 | 77561 | 40 |
|  | **Male** | 15 | 62 | 560 | 63 | 8401 | 60 | 25662 | 60 | 23319 | 58 | 20706 | 60 | 17983 | 60 | 18241 | 61 | 114889 | 60 |
| **Age group** | **≤20** | 3 | 13 | 129 | 15 | 1776 | 13 | 4523 | 11 | 5373 | 13 | 4418 | 13 | 2789 | 9 | 2437 | 8 | 21448 | 11 |
|  | **21-40** | 9 | 38 | 400 | 45 | 6045 | 43 | 17051 | 40 | 15445 | 38 | 13504 | 39 | 11164 | 38 | 11017 | 37 | 74635 | 39 |
|  | **41-60** | 4 | 17 | 265 | 30 | 4639 | 33 | 15055 | 35 | 13436 | 33 | 11777 | 34 | 10563 | 35 | 10877 | 36 | 66616 | 35 |
|  | **61-80** | 7 | 29 | 79 | 9 | 1346 | 10 | 5873 | 14 | 5414 | 13 | 4627 | 13 | 4676 | 16 | 4971 | 17 | 26993 | 14 |
|  | **>80** | 1 | 4 | 9 | 1 | 115 | 1 | 542 | 1 | 543 | 1 | 473 | 1 | 574 | 2 | 510 | 2 | 2758 | 1 |
| **Total cases** | | **24** | **100** | **882** | **100** | **13921** | **100** | **43044** | **100** | **40211** | **100** | **34799** | **100** | **29766** | **100** | **29812** | **100** | **192450** | **100** |
| **COVID-19 deaths** | |  |  |  |  |  |  |  |  |  |  |  |  |  |  |  |  |  |  |
| **Sex** | **Male** | 1 | 100 | 16 | 73 | 229 | 65 | 818 | 72 | 522 | 68 | 383 | 68 | 281 | 70 | 227 | 71 | 2452 | 69 |
|  | **Female** | 0 | 0 | 6 | 27 | 122 | 35 | 323 | 28 | 250 | 32 | 184 | 32 | 119 | 30 | 94 | 29 | 1091 | 31 |
| **Age group** | **≤20** | 0 | 0 | 0 | 0 | 1 | 0 | 3 | 0 | 4 | 1 | 2 | 0 | 0 | 0 | 1 | 0 | 11 | 0 |
|  | **21-40** | 0 | 0 | 2 | 9 | 24 | 7 | 60 | 5 | 43 | 6 | 19 | 4 | 11 | 3 | 7 | 2 | 166 | 5 |
|  | **41-60** | 0 | 0 | 11 | 50 | 139 | 40 | 374 | 33 | 210 | 27 | 148 | 26 | 107 | 27 | 62 | 19 | 1051 | 30 |
|  | **61-80** | 1 | 100 | 7 | 32 | 164 | 47 | 574 | 50 | 409 | 53 | 298 | 53 | 209 | 52 | 190 | 59 | 1852 | 52 |
|  | **>80** | 0 | 0 | 2 | 9 | 22 | 6 | 128 | 11 | 102 | 13 | 89 | 16 | 59 | 15 | 61 | 19 | 463 | 13 |
| **Total deaths** | | **1** | **100** | **22** | **100** | **350** | **100** | **1139** | **100** | **768** | **100** | **556** | **100** | **386** | **100** | **321** | **100** | **3543** | **100** |
